# Agreement of parent- and child-reported wheeze: are they associated with FeNO and lung function?

**DOI:** 10.1101/2021.03.19.21253977

**Authors:** Rebeca Mozun, Cristina Ardura-Garcia, Eva S. L. Pedersen, Myrofora Goutaki, Jakob Usemann, Florian Singer, Philipp Latzin, Alexander Moeller, Claudia E. Kuehni

## Abstract

In epidemiological studies, childhood asthma is usually assessed with questionnaires directed at parents or children, and these may give different answers. We studied how well parents and children agreed when asked to report symptoms of wheeze and investigated whose answers were closer to measurable traits of asthma.

*LuftiBus in the school* is a cross-sectional survey of respiratory health among Swiss schoolchildren aged 6-17 years. We applied questionnaires to parents and children asking about wheeze and exertional wheeze in the past year. We assessed agreement between parent-child answers with Cohen’s kappa (k), and associations of answers from children and parents with physiological measurements (i.e. FeNO and FEV1/FVC), using quantile regression.

We received questionnaires from 3079 children and their parents. Agreement was poor for reported wheeze (k=0.37) and exertional wheeze (k=0.36). Median FeNO varied when wheeze was reported by children (19 ppb, IQR: 9-44), parents (22 ppb, IQR: 12-46), both (31 ppb, IQR: 16-55) or neither (11 ppb, IQR: 7-19). Median absolute FEV1/FVC was the same when wheeze was reported by children (84%, IQR: 78-89) and by parents (84%, IQR: 78-89), lower when reported by both (82%, IQR: 78-87) and higher when reported by neither (87%, IQR: 82-91). For exertional wheeze findings were similar. Results did not differ by age or sex.

Our findings suggest that surveying both parents and children and combining their responses can help us to better identify children with measurable asthma traits.

**Take home message:** There is poor agreement between schoolchildren and their parents when reporting current wheeze. However, wheeze correlates best with lung function and FeNO when reported by both children and parents.

## Introduction

Asthma is a complex disease and physicians use a combination of symptoms, lung function and airway inflammation to diagnose it [1-3]. In large epidemiological surveys, however, researchers usually rely on reported wheeze to assess prevalence, risk factors and prognosis of asthma [4-6].

Reports of wheeze differ between parents and children and it is unclear whose reports are a more accurate indication of asthma measured by physiological traits such as airflow limitation or airway inflammation [3, 7]. Knowledge on this would help to decide whether researchers should question parents or children, or both. The latter approach might be more informative but does increase complexity and costs of the study. Three previous studies compared answers from parents and children to measurable asthma traits [8-10]. Two were done in asthma outpatient clinics and used composite symptom scores [8, 9]. One was a population-based study [10]. All three studies used spirometry to assess airflow limitation and compared spirometry results to reported wheeze, but none combined answers from parents and children to consider their agreement. We compared wheeze reported by children, by parents and by both, to two asthma traits which measure different aspects of this complex disease: airflow limitation assessed by spirometry and eosinophilic airway inflammation assessed by Fractional exhaled Nitric Oxide, FeNO. We measured agreement between parents’ and children’s reports of wheeze and exertional wheeze, identified determinants of agreement, and examined the association with lung function and FeNO.

## Methods

### Study design and setting

*LuftiBus in the* school (LUIS) is a cross-sectional study performed between 2013 and 2016 in schools of the canton of Zurich, Switzerland. Recruitment methods and procedures have been described [11]. In short, all schools in the canton were invited to participate. If the head teacher agreed, trained lung function technicians visited the school with a mobile lung function lab in a bus [12]. Parents completed a detailed questionnaire at home. Children were interviewed and underwent lung function tests at school. Technicians were not aware of the answers to the parental questionnaire. For this analysis, we included all children with consent to participate for whom we had both a child’s questionnaire and a parent completed questionnaire. The ethics committee of the canton of Zurich approved the study (KEK-ZH-Nr: 2014-0491).

### Questionnaire design and definitions

The parental questionnaire was printed and completed at home. It asked about respiratory diagnoses, symptoms and their trigger factors, medication, parental history of atopic diseases, family and household characteristics, child’s country of birth and parents’ countries of origin. Questions came from the International Study of Asthma and Allergies in Childhood (ISAAC) and the Leicester Respiratory Cohort studies questionnaires [13, 14]. We asked who completed the parental questionnaire (mother or father) and whether the child helped to complete it. The questionnaire for children was short and completed online by study technicians who interviewed the children at school [15]. Children were asked key questions on respiratory symptoms and triggers of wheeze. Questionnaires to both children and parents included a written explanation of the term wheeze. The wording in the child and parental questionnaires was almost identical with slight simplifications in the child’s version (Table S1) [16].

### FeNO and spirometry

FeNO was measured before spirometry with a fast response chemiluminescence analyser CLD 88, Eco Medics AG, Duernten, Switzerland, and expressed as parts per billion (ppb). For spirometry we used Masterlab, Jaeger, Würzburg, Germany, according to ERS/ATS standards [17]. Our main outcomes were the ratio of forced expiratory volume in one second and forced vital capacity (FEV1/FVC) in absolute percent, z-scores of FEV1 and z-scores of forced expiratory flow between 25% and 75% of the FVC (FEF25-75) using Global Lung Initiative (GLI) references [18]. Quality criteria of flow-volume curves were assessed post hoc and only valid tests were included in the analysis [11].

### Statistical analysis

We assessed agreement between answers from parents and children to questions on wheeze and on exertional wheeze by cross-tabulation and calculated unweighted Cohen’s kappa to adjust for agreement by chance [19, 20]. We interpreted kappa as: 0-0.20=none, 0.21-0.39=poor, 0.40-0.59=weak, 0.60-0.79=moderate, 0.80-0.89=strong, 0.90-1=almost perfect agreement [19]. Questions about triggers of wheeze had also a “don’t know” answer category, which we recoded as “no” to simplify the analysis.

We then studied the determinants of agreement between answers from parents and children using multinomial logistic regression. The models had three possible outcomes: parents and children both answered “yes” (agreed for yes), parents and children both answered “no” (agreed for no), and parents and children disagreed (reference). We chose possible determinants a priori based on the literature [16, 21]: age, sex, child’s country of birth, parents’ countries of origin, number of children in the household, neighbourhood socioeconomic position index (Swiss SEP) [11, 22, 23], urbanisation degree of their place of residence [24], parental history of asthma, and the person completing the parental questionnaire.

Lastly, we studied the associations of parent-child reported wheeze and exertional wheeze with FeNO and lung function. For the analysis, we used three scenarios, which reflect hypothetical studies where questionnaires were sent only to children (scenario A), only to parents (scenario B) or to both (scenario C). Main outcomes were FeNO as a measure of airway inflammation and FEV1/FVC as a sensitive measure of airflow limitation. We reported median values for FeNO and FEV1/FVC, as they were not normally distributed. We calculated median differences in FeNO and FEV1/FVC between children with wheeze and children without wheeze for each of the three scenarios using quantile regressions. Additional lung function outcomes were z-scores of FEV1 and FEF25-75. There, we used linear regression to calculate mean differences between children with and without wheeze in all three scenarios. Age and sex were included as confounders in all models. In a sensitivity analysis we also adjusted for whether the child had helped the parents in completing their questionnaire.

We assessed differences between the scenarios by calculating the difference in median FeNO and FEV1/FVC and in mean z-scores of FEV1 and FEF 25-75 in children with wheeze between scenario A (child-reported) and B (parent-reported), scenario A and C (parent- and child-reported), and scenario B and C. We calculated 95% confidence intervals using the bootstrap method with 500 repetitions and considered there was a statistically significant difference between scenarios when the 95% confidence intervals excluded 0. We reported these results also stratified by age, in children aged less than 10 years and 10 years or more, to compare our findings with the literature [8-10]. We used the software STATA (Version 16.1, StataCorp, TX) for statistical analysis, and followed STROBE reporting guidelines [25].

## Results

### Study population

490 schools from the canton of Zurich were invited and 37 participated. 3870 children aged 6-17 years took part. For 3079 we had questionnaires with information on wheeze and exertional wheeze from both parents and children (Table 1). FeNO results were available for 2762 children (median 12ppb, interquartile range [IQR] 7-21) and FVC measurements for 2217 (median 87%, IQR 82-91). Most questionnaires were completed by the mothers (n=1765, 57%). 667 children (22%) helped their parents to complete the questionnaire. Parents of 425 children (14%) had a history of asthma.

**Table 1:**
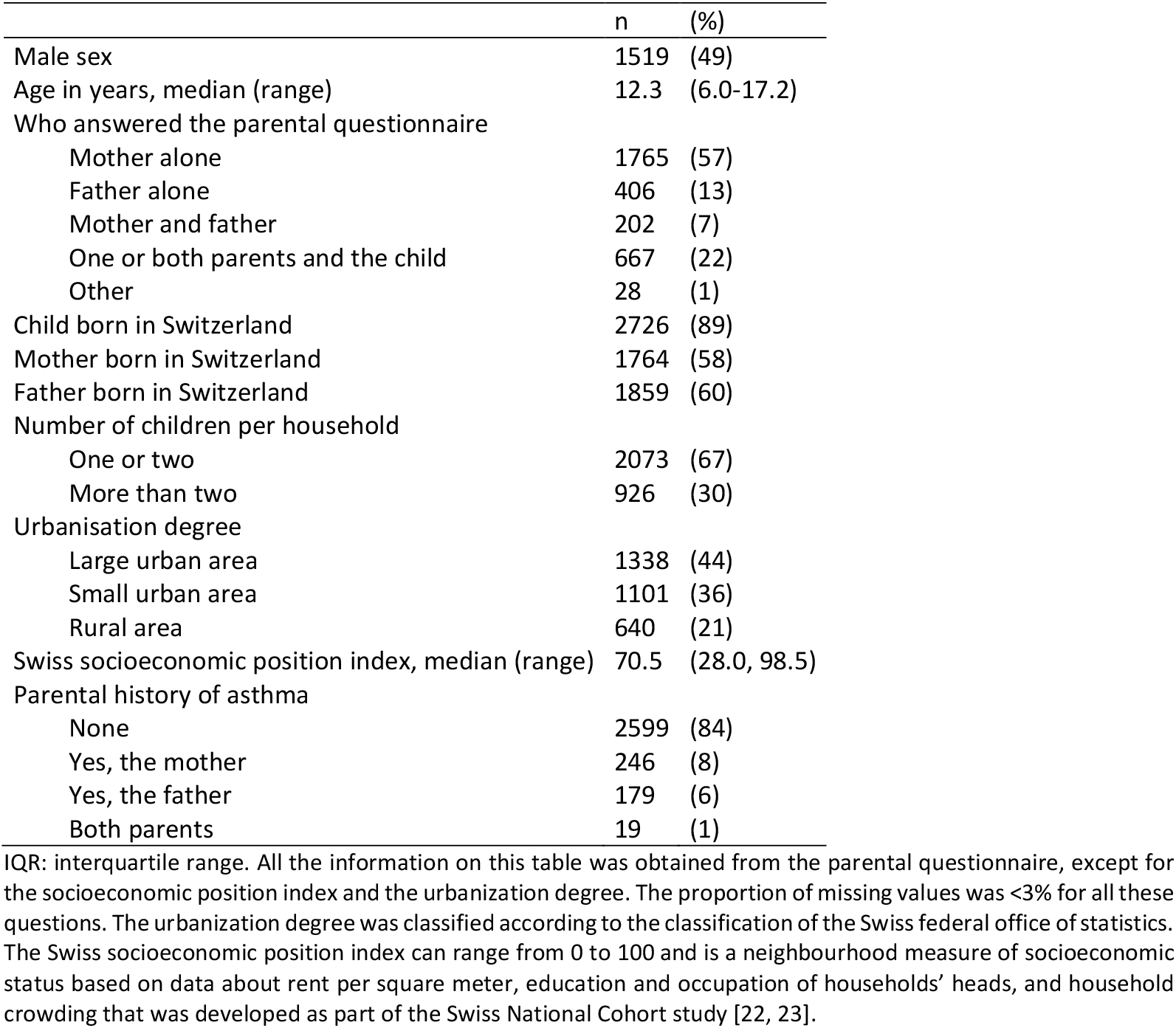
Sociodemographic characteristics of schoolchildren in the LUIS study (N=3079).

### Prevalence of wheeze and agreement between parents and children

The prevalence of wheeze was comparable, independent of whether information arose from parents or from children (Figure 1, left). However, the group of children who reported wheeze was not the same as the group whose parents reported wheeze, although there was an overlap (Figure 1, right). The occurrence of wheeze in the past 12 months was reported by 273 (9%) children and 236 (8%) parents (p value for difference in proportions 0.090). Only in 108 (4%) wheeze was reported by both parents and children. Exertional wheeze in the past 12 months was reported by 12% of children (n=369) and 8% of parents (n=234; p value <0.001), and by both in only 4% (n=127). Using kappa (k) statistics, we found poor agreement between parents’ and children’s reports for wheeze (k=0.37) and exertional wheeze (k=0.36) (Figure 1). Agreement was poor for reported triggers of wheeze, lowest for colds or infections (k=0.12) and highest for pets (k=0.40).

**Figure 1:**
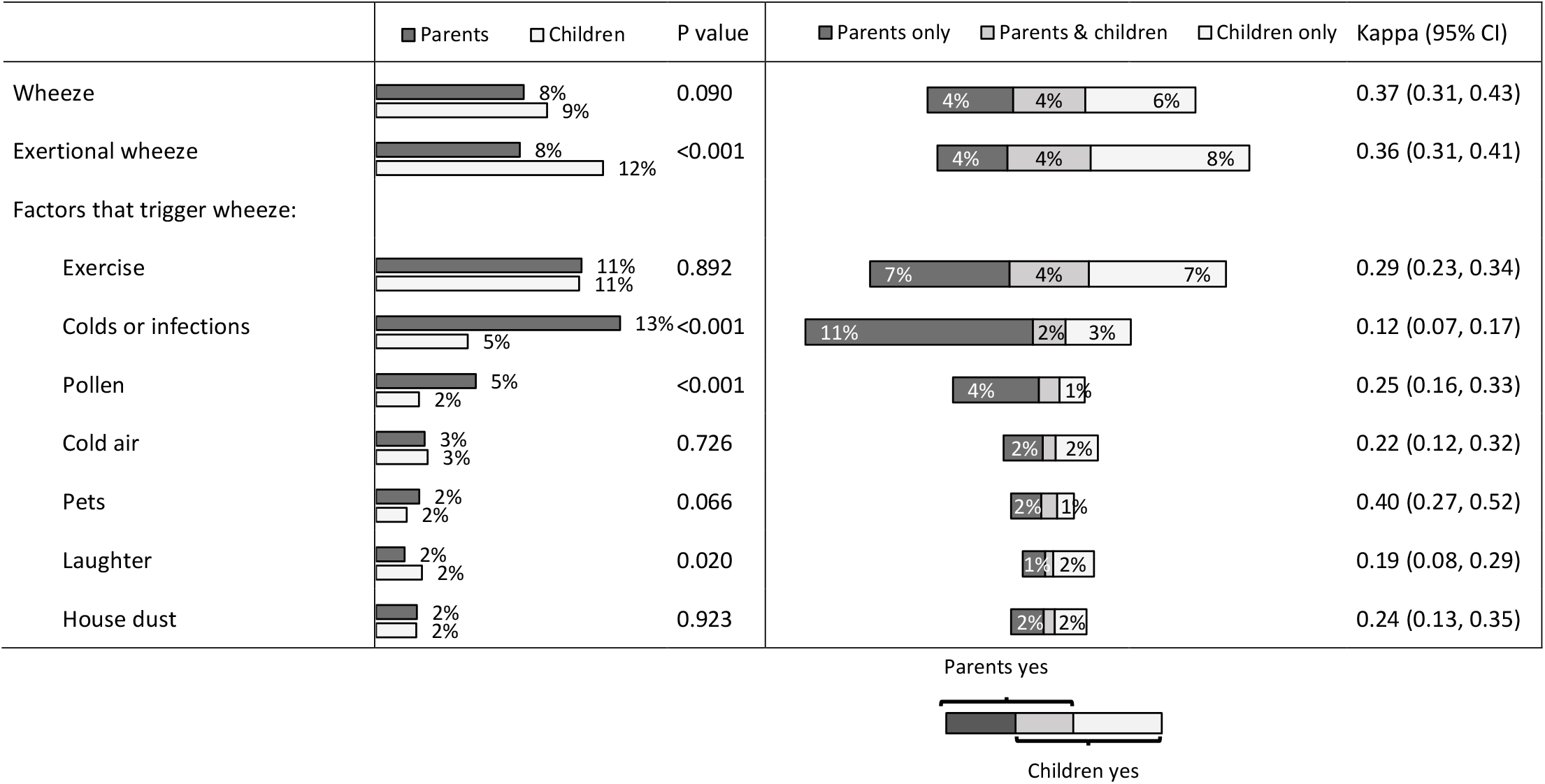
Parent and child reported prevalence of wheeze in the past 12 months, and proportion of agreement between their answers (N=3079). Differences in total percentages between both columns are due to rounding. P value for a difference between two proportions (parent vs child reported symptoms). Kappa = 1 means perfect agreement. Kappa = 0 means agreement due to chance. Online table S2 shows the exact numbers and prevalence confidence intervals for this figure.

### Determinants of agreement

Children’s age, parental history of asthma and whether the child helped to complete the parental questionnaire determined the agreement between parents’ and children’s reported wheeze, after adjustment for sex, socioeconomic and family characteristics (Figure S1). Older children agreed less with their parents when answering “no” to exertional wheeze (age 10-13 vs 6-9 years: relative risk ratio [RRR]: 0.5, 95% confidence interval [CI]: 0.4-0.7; age 14-17 vs 6-9 years: RRR: 0.6, 95% CI: 0.4-0.9) (Figure S1, right panel). We found weak evidence that boys agreed less often than girls with their parents when reporting wheeze (RRR: 0.7, 95% CI: 0.5-1.2) and exertional wheeze (RRR: 0.7, 95% CI: 0.5-1.1) and also when answering “no” to wheeze (RRR: 0.8, 95% CI: 0.6-1.1) or exertional wheeze (RRR: 0.9, 95% CI: 0.7-1.2). Families with more than two siblings agreed more on wheeze (RRR: 1.6, 95%CI 1.0-2.5), but not on exertional wheeze. We found no influence on agreement of socioeconomic position, country of birth of the child and of the parents. If parents had a history of asthma, parents and children were more likely to agree on wheeze (RRR: 1.5, 95% CI: 0.9-2.4) or exertional wheeze (RRR: 2.0, 95% CI: 1.2-3.1), but less likely to agree on answering “no” to wheeze (RRR: 0.4, 95% CI: 0.3-0.5) or exertional wheeze (RRR: 0.5, 95% CI: 0.4-0.7). When the child helped to complete the parental questionnaire, parents and children were more likely to agree on reporting wheeze (RRR: 2.4, 95% CI: 1.4-4.2) or exertional wheeze (RRR: 1.8, 95% CI: 1.1-2.8).

### Association between reported wheeze and FeNO and lung function

FeNO was higher in children who reported wheeze (19ppb, interquartile range [IQR]: 9-44) than in those who did not (12ppb, IQR: 7-20) (Figure 2, A; Table S3). When considering parental reports (Figure 2, B), median FeNO was 22ppb (IQR: 12-46) for children with wheeze compared to 12ppb (IQR: 7-20) for those without. When we used both sources of information (Figure 2, C), FeNO was highest when parents and children both reported wheeze (31ppb, IQR: 16-55), intermediate when only one of them reported wheeze (17ppb, IQR: 8-34) and lowest when neither parents nor children reported wheeze (11ppb, IQR: 7-19). For exertional wheeze findings were similar (Figure 2, right panel). We estimated the difference in median FeNO between children with and without wheeze as reported by children or parents or both, adjusted for age and sex in a regression model. The difference was largest when both reported wheeze (21ppb, 95% CI: 18-23) or exertional wheeze (13ppb, 95% CI: 11-16) (Table S3). We also compared differences in FeNO between scenarios (Table S4). FeNO was similar between children with child-reported wheeze and children with parent-reported wheeze (scenarios A-B, p value 0.236), but was higher when wheeze was reported by both than by children (scenarios A-C, p value 0.003) or parents (scenarios B-C, p value 0.020). When we stratified by age, older children had higher FeNO when wheeze was reported by parents than by children (scenarios A-B), with no differences among younger children (Table S5). Lung function was lower in children with wheeze than in those without wheeze when reported by both children and parents. Results followed the same pattern as for FeNO. There was a difference in FEV1/FVC between groups of children with and without wheeze (Figure 3). When children reported wheeze, median FEV1/FVC was 84% (IQR 78-89) compared to 87% (IQR: 82-91) when they did not (scenario A). We found the same when using answers from parents (scenario B). When we combined answers from parents and children (scenario C), median FEV1/FVC was lowest when both parents and children reported wheeze (82%, IQR: 78-87), compared to when only one of them reported wheeze (85%, IQR: 79-89), and when neither reported wheeze (87%, IQR: 82-91). Findings for exertional wheeze were similar. We obtained similar results when we adjusted for age and sex in regression analyses, and when we used FEV1 or FEF 25-75 z-scores as outcomes instead of FEV1/FVC (Table S3). When we compared results between scenarios, we found similar lung function results in children with wheeze between scenarios A and B (p value for FEV1/FVC: 0.964), but lung function was lower in scenario C than A (p value 0.045) or B (p value 0.030) (Table S4). The associations with lung function did not vary by age (Table S5).

**Figure 2:**
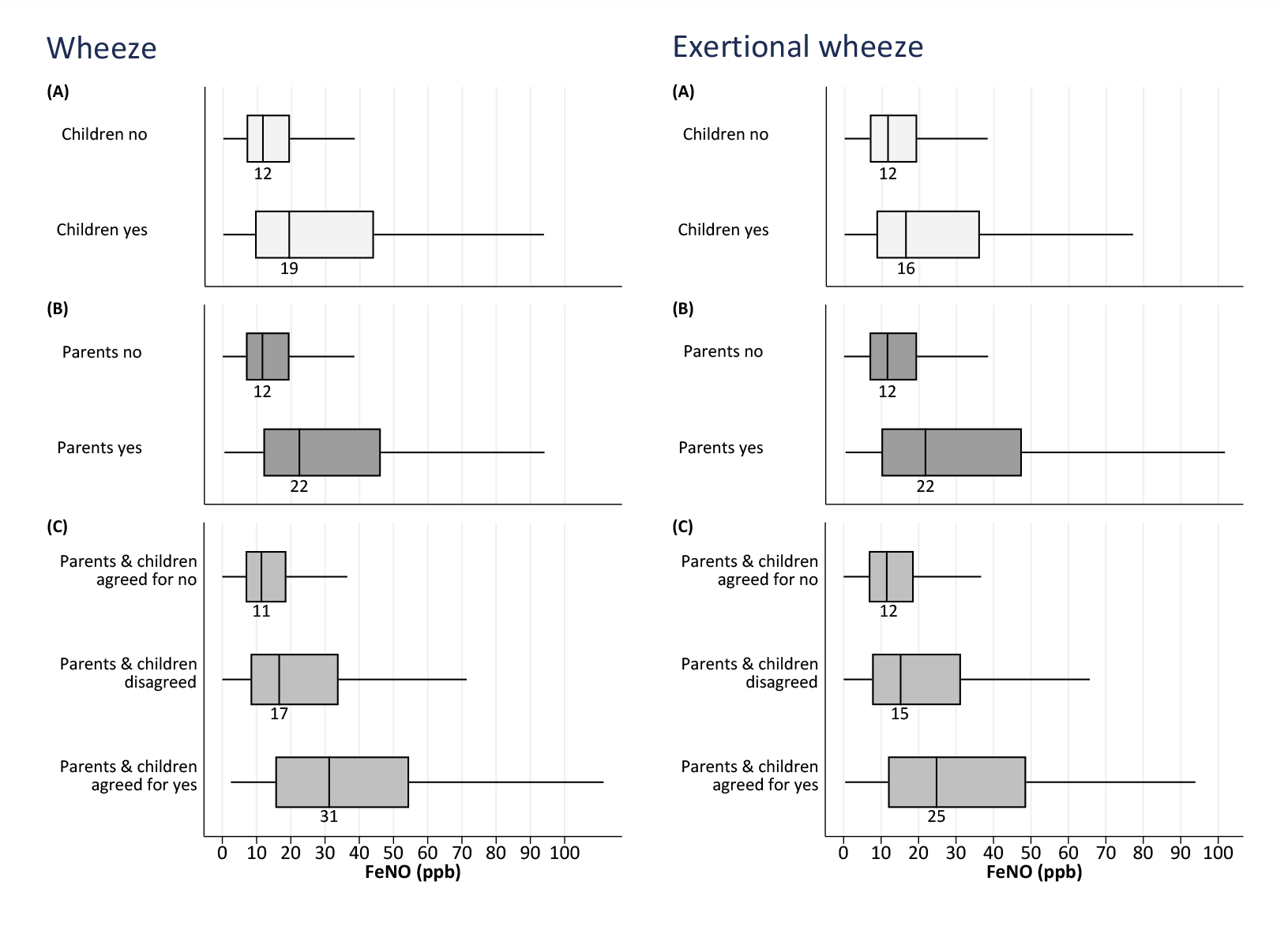
Distribution of FeNO in schoolchildren with and without wheeze as reported by (A) children, (B) parents, (C) either parents or children, and both parents and children (N=2762). FeNO = Fractional exhaled Nitric Oxide, measured in parts per billion (ppb). Wheeze and exertional wheeze refer to the past 12 months. The left vertical lines of the boxes represent 25^th^ percentiles, middle lines and data labels show the median, and right lines show the 75^th^ percentiles of FeNO values. Outliers not displayed.

**Figure 3:**
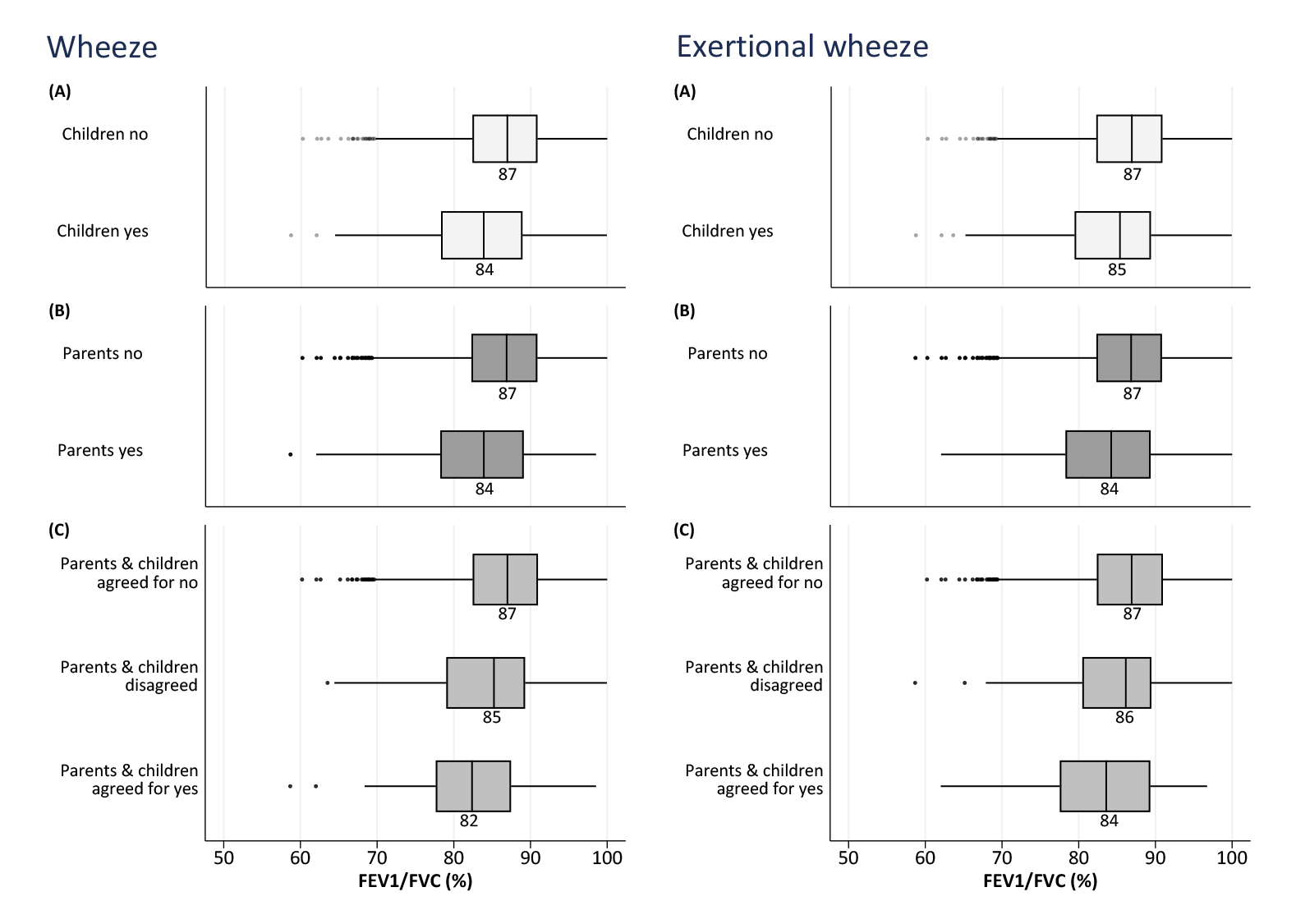
Distribution of FEV1/FVC in schoolchildren with and without wheeze as reported by (A) children, (B) parents, (C) either parents or children, and both parents and children (N=2217). FEV1/FVC = Forced Expiratory Volume in one second over Forced Vital Capacity, measured in percentage. Wheeze and exertional wheeze refer to the past 12 months. The left vertical lines of the boxes represent 25th percentiles, middle lines and data labels show the median, and right lines the 75th percentiles of FEV1/FVC.

Additional adjustment for the child’s help in completing questionnaires did not change the results for any model (data not shown).

## Discussion

This study found that parents and children agree poorly when reporting wheeze. When comparing parents’ and children’s reporting of wheeze with measurable traits of asthma, we found that lung function and FeNO differed depending on who reported wheeze. FeNO levels were highest and airflow was more limited in children for whom both children and parents reported wheeze.

### Comparison with other studies

Few studies compared parent and child reported wheeze. Some found the same prevalence of wheeze when they questioned parents or children [26, 27], while others observed a higher prevalence when questioning adolescents rather than their parents (Table 2) [21, 28, 29]. Hedman described that exertional wheeze was reported by 14% of teenagers and 8% of their parents [27]. Children may be more aware of mild symptoms during exercise than their parents, which may also explain why in our study child-reported exertional wheeze was less strongly associated with measurable asthma traits than parent-reported.

**Table 2:** Prevalence of parent- and child-reported wheeze and kappa estimates for chance-adjusted agreement between parents and children in different studies.

| Study | LUIS | Braun-Fahländer [21] | Hedman [27] | Renzoni [28] | Mallol [26] | Decker [29] |
| --- | --- | --- | --- | --- | --- | --- |
| Country of the study | CH | CH | SE | IT | CL | US |
| Study sample size (N) | 3113 | 1374 | 294 | 21068 | 3178 | 230 |
| Age of study population (years) | 5-17 | 13-15 | 13-14 | 13-14 | 13-14 | 10-12 |
| Respiratory symptoms: |  |  |  |  |  |  |
| Wheeze |  |  |  |  |  |  |
| Child-reported prevalence | 9% | 11% | 8% | 10% | 11% | 11% |
| Parent-reported prevalence | 8% | 7% | 7% | 5% | 10% | 6% |
| Kappa | 0.37 | 0.48 | 0.61 | 0.44 | 0.29 | 0.18 |
| Exertional wheeze |  |  |  |  |  |  |
| Child-reported prevalence | 12% | 20% | 14% | - | 26% | 25% |
| Parent-reported prevalence | 8% | 8% | 8% | - | 13% | 12% |
| Kappa | 0.36 | 0.39 | 0.44 | - | 0.21 | 0.38 |

Wheeze and exertional wheeze refer to the past 12 months. Abbreviations: CH= Switzerland, SE= Sweden, IT= Italy, CL= Chile, US= United States.

In our study, the agreement between answers from parents and children was poor both for current wheeze (kappa 0.37) and exertional wheeze (kappa 0.36). Other studies found similarly weak agreement rates with kappa estimates for parent-child agreement in the range of 0.18-0.61 for current wheeze and 0.21-0.44 for exertional wheeze (Table 2) [21, 26-28, 30]. A reason could be that the understanding of the term wheeze by parents and children is limited [16, 31]. Definitions of wheeze from parents attending respiratory clinics often differ from definitions given in surveys [31]. Also a population-based study found that parental understanding of wheeze was moderate, but better for children with severe asthma, and those whose mothers had asthma, and worse for children of ethnic minorities and those living in deprived areas [16]. In line with our findings, Braun-Fahrländer described better parent-child agreement for girls, when children helped to complete questionnaires and when parents had a history of asthma [21]. The latter could be due to better recognition of wheeze by parents, increased awareness in children, or higher prevalence of wheeze, when parents had asthma.

Few studies have compared parent- and child-reported symptoms with measurable traits of asthma [8-10]. Horak compared lung function with parent- and child-reported asthma symptoms using symptom scores among 90 school aged children with asthma [8]. They found that among children younger than 10 years with low lung function, parents reported more symptoms (higher symptom scores) than their children. Guyat assessed symptoms in 52 children with asthma aged 7-17 years and found that in children below 10 years of age, parent-reported symptom scores correlated better with lung function than symptom scores reported by the children, whereas the opposite was found for the children older than 10 years [9]. Yu compared lung function in 1963 schoolchildren aged 8-12 years with reports from parents and children [10]. Children aged less than 10 years were more likely to have low FEV1/FVC if wheeze was reported by parents (OR 3.1) compared to children (OR 1.5), but no difference was found when children were older than 10 years. We found that the association with lung function and wheeze was similar for parent-and child-reported wheeze. FeNO was higher when wheeze was reported by parents than by children for children aged 10 or more, but not for younger children. Previous studies did not investigate the usefulness of combining answers from parents and children to assess agreement. In our study, when parents and children both reported wheeze or exertional wheeze, children had the highest FeNO and worst lung function, indicative of asthma.

### Strengths and limitations

This is the first study that investigated the association of parent and child reported wheeze with two different traits related to asthma, FeNO and lung function. In contrast to others, we did not only assess answers from parents and children separately (scenarios A and B) but also combined (scenario C). Further strengths are our large sample size and comprehensive evaluation of lung function. One limitation of our study is that parents completed the questionnaire at home while children were interviewed in the bus at school by a study technician. This may have helped children to understand the term “wheeze” so we cannot extrapolate our results to a situation where children answer printed questionnaires. Questionnaires to parents and children contained explanations what we mean by wheeze. Ideally, a diagnosis of asthma should be done based on the clinical history and at least two physiological measurements [32]. This was not possible in our study. However, we compared the responses for wheezing and exertional wheezing with the two most widely recommended measurable traits for asthma: airflow limitation and airway inflammation. We acknowledge the fact that children with current asthma may have normal lung function and atopic children may show elevated FeNO without asthma. However, we believe that this should not have significantly biased the comparison between the three scenarios, since all scenarios include the same children.

### Implications and conclusion

Epidemiological studies commonly rely on reported wheeze as a proxy measure for asthma. Our study raises awareness of the fact that parents and children do not report wheeze consistently. Our findings suggest that using reports from both parents and children may bring us closer to measurable traits of asthma. This highlights the importance of asking both parents and children about wheeze in epidemiological studies but may also be valid for clinical practice. Researchers may be able to distinguish children with a greater likelihood or severity of asthma when both, parents and children, report wheeze. Similarly, asthma is unlikely when both independently do not report wheeze. Our results suggest that future epidemiological studies on childhood asthma should whenever possible address both, parents and children.

## Data Availability

Study collaborators and other researchers can obtain datasets for analysis if a detailed concept sheet is presented for the planned analyses and approved by the principal investigators (AM, PL and CK).

## Acknowledgements

We thank the staff from the schools, children, and their families for taking part in the study, and fieldworkers of the LUIS study for their technical support during the study. We thank Ben Spycher and Marcel Zwahlen (Institute of Social and Preventive Medicine, University of Bern, Switzerland) for their statistical advice. We thank Daria O. Berger (Institute of Social and Preventive Medicine, University of Bern) for her English language editing contributions to the manuscript. We thank Johanna M. Kurz, Andras Soti, Marc-Alexander Oestreich, Corin Willers (Paediatric Respiratory Medicine, Children’s University Hospital of Bern, University of Bern), Léonie Hüsler, Eugénie Collaud and Carmen C. M. de Jong (Institute of Social and Preventive Medicine, University of Bern) for their help in the assessment of the quality of the spirometry flow-volume curves.

## The LuftiBus in the school study group

The study PIs Prof. Alexander Moeller, Prof. Claudia E. Kuehni and Prof. Philipp Latzin conceptualised and designed the study. Alexander Moeller supervised data collection and was responsible for the overall conduct of the study. Philipp Latzin was responsible for the lung function measurements. Claudia E. Kuehni was responsible for the questionnaire design and data analysis. Rebeca Mozun was responsible for data management and coordination. Florian Singer and Johanna Kurz (Paediatric Respiratory Medicine, Children’s University Hospital of Bern, University of Bern, Switzerland) are responsible for management and analysis of DTG-SBW data. Myrofora Goutaki, Eva S.L. Pedersen and Cristina Ardura-Garcia support statistical analysis. Jakob Usemann and Kees de Hoogh (Swiss Tropical and Public Health Institute, Basel, Switzerland) are responsible for the air pollution data.

## Authors’ contributions

Claudia E. Kuehni, Alexander Moeller and Phillip Latzin conceptualised and designed the study. Alexander Moeller supervised data collection. Rebeca Mozun analysed the data and drafted the manuscript. Cristina Ardura-Garcia and Eva S.L. Pedersen supported the statistical analysis. All authors gave input for interpretation of the data. All authors critically revised and approved the manuscript.

