## Supplement for "Agreement of parent- and child-reported wheeze: are they associated with FeNO and lung function?"

### Online supplementary material

**Table of contents:**

**Table S1:** Original questions in German (A) and English (B) language from the parental questionnaire and children's questionnaire used in the LuftiBus in the school (LUIS) study……………………………………..2

**Table S2:** Prevalence of parent- and child-reported wheeze and proportion of children with wheeze by groups of agreement in answers from parents and children…………………………………………………………3

**Table S3:** Median difference in FeNO and FEV1/FVC and mean difference in FEV1 and FEF25-75 in schoolchildren with wheeze when reported by (A) children, (B) parents, and (C) parents and children, compared to children with no reported wheeze……………………………………………………………………………….4

**Table S4:** Difference in median FeNO and FEV1/FVC and mean FEV1 and FEF25-75 in schoolchildren with wheeze between scenario A (child-reported) and B (parent-reported), scenario A and C (parent- and child-reported), and scenario B and C………………………………………………………………………………….5

**Table S5:** Difference in median FeNO and FEV1/FVC in schoolchildren with wheeze when reported by children compared to reported by parents stratified by age groups…………………………………………………6

**Figure S1:** Characteristics of parent-child pairs who agreed for yes (1) and for no (2), compared to parent-child pairs who disagreed in their answers……………………………………………………………………………..7

**Table S1:** Original questions in German (A) and English (B) language from the parental questionnaire and children's questionnaire used in the LuftiBus in the school (LUIS) study.

| | Parental questionnaire | Children's questionnaire |
|---|---|---|
| **A) German** | | |
| Explanation | Die Fragen im nächsten Teil beziehen sich auf pfeifende oder keuchende Atmung. Darunter verstehen wir Geräusche, die beim Atmen (vor allem beim Ausatmen) aus dem Brustkorb kommen, nicht aus der Nase. | Jetzt fragen wir dich nach der Atmung in den letzten 12 Monaten. Es gibt Kinder, bei denen es manchmal beim Atmen pfeifende oder keuchende Geräusche im Brustkorb gibt. |
| Wheeze | Hatte Ihr Kind in den letzten 12 Monaten beim Atmen pfeifende oder keuchende Geräusche im Brustkorb?<br>*Ja; Nein* | Hattest du in den letzten 12 Monaten beim Atmen manchmal pfeifende oder keuchende Geräusche im Brustkorb?<br>*Nein; Ja* |
| Exertional wheeze | Hatte Ihr Kind in den letzten 12 Monaten pfeifende oder keuchende Geräusche im Brustkorb während oder nach körperlicher Anstrengung?<br>*Ja; Nein* | Hattest du in den letzten 12 Monaten jemals pfeifende oder keuchende Geräusche im Brustkorb während oder nach körperlicher Anstrengung (Rennen, Velofahren)?<br>*Nein; Ja* |
| Wheeze triggers | Welche der folgenden Situationen hat bei Ihrem Kind in den letzten 12 Monaten Husten oder pfeifende/keuchende Atmung ausgelöst? (Bitte kreuzen Sie alle zutreffenden Antworten an.)<br>Pfeifende/keuchende Atmung<br>Anstrengung (Rennen, Velofahren, etc.)<br>*Ja; Nein; Weiss nicht*<br>Hausstaub<br>*Ja; Nein; Weiss nicht*<br>Bluütenstaub (Gras, Heu, Blumen, Bäume)<br>*Ja; Nein; Weiss nicht*<br>Kontakt mit Tieren<br>*Ja; Nein; Weiss nicht*<br>Kalte Luft oder Nebel<br>*Ja; Nein; Weiss nicht*<br>Lautes Lachen<br>*Ja; Nein; Weiss nicht*<br>Erkältungen<br>*Ja; Nein; Weiss nicht* | Welche der folgenden Situationen hat bei dir in den letzten 12 Monaten Husten oder pfeifende/keuchende Atmung oder beides ausgelöst?<br>Pfeifende/keuchende Atmung /Atemnot<br>Anstrengung (Rennen, Velo, etc.)<br>*Ja; Nein; Weiss nicht*<br>Hausstaub<br>*Ja; Nein; Weiss nicht*<br>Bluütenstaub (Gras, Heu, Blumen, Bäume)<br>*Ja; Nein; Weiss nicht*<br>Kontakt mit Tieren<br>*Ja; Nein; Weiss nicht*<br>Kalte Luft oder Nebel<br>*Ja; Nein; Weiss nicht*<br>Lautes Lachen<br>*Ja; Nein; Weiss nicht*<br>Erkältungen<br>*Ja; Nein; Weiss nicht* |
| **B) English** | | |
| Explanation | Questions on wheezing. By "wheeze" we mean breathing that makes a high-pitched sound whistling or squeaking sound from the chest not the throat. | By "wheezing" we mean breathing that makes a high-pitched whistling or squeaking sound from the chest, not the throat. |
| Wheeze | Has your child had wheezing or whistling in the chest in the last 12 months?<br>*Yes; No* | Have you had wheezing or whistling in the chest in the last 12 months?<br>*No; Yes* |
| Exertional wheeze | In the past 12 months, has your child's chest sounded wheezy during or after exercise?<br>*Yes; No* | In the past 12 months, has your chest sounded wheezy during or after exercise? (racing, cycling)?<br>*No; Yes* |
| Wheeze triggers | In the past 12 months, did the following things cause coughing or wheezing in your child?<br>(Please tick all applicable answers)<br>Wheeze<br>Exercise (running, biking, etc.)<br>*Yes; No; Don't know*<br>House dust<br>*Yes; No; Don't know*<br>Pollen (grass, hay, trees)<br>*Yes; No; Don't Know*<br>Contact with animals<br>*Yes; No; Don't know*<br>Cold temperatures or fog<br>*Yes; No; Don't know*<br>Intense laughing<br>*Yes; No; Don't know*<br>Colds<br>*Yes; No; Don't Know* | In the past 12 months, did the following things cause you coughing or wheezing?<br>(Please tick all applicable answers)<br>Wheeze<br>Exercise (running, biking, etc.)<br>*Yes; No; Don't know*<br>House dust<br>*Yes; No; Don't know*<br>Pollen (grass, hay, trees)<br>*Yes; No; Don't Know*<br>Contact with animals<br>*Yes; No; Don't know*<br>Cold temperatures or fog<br>*Yes; No; Don't know*<br>Intense laughing<br>*Yes; No; Don't know*<br>Colds<br>*Yes; No; Don't Know* |

**Table S2:** Prevalence of parent- and child-reported wheeze and proportion of children with wheeze by groups of agreement in answers from parents and children.

| | | (A) | | (B) | | (C) | | | | | | | |
|---|---|---|---|---|---|---|---|---|---|---|---|---|---|
| | N | Children | | Parents | | Parents & children agreed for yes | | Parents only | | Children only | | Parents & children agreed for no | |
| | | n | % [95% CI] | n | % [95% CI] | n | % | n | % | n | % | n | % |
| Wheeze | 2982 | 273 | 9% [8-10] | 236 | 8% [7-9] | 108 | 4% | 128 | 4% | 165 | 6% | 2581 | 87% |
| Exertional wheeze | 3037 | 369 | 12% [11-13] | 234 | 8% [7-9] | 127 | 4% | 107 | 4% | 242 | 8% | 2561 | 84% |
| Factors that trigger wheeze | | | | | | | | | | | | | |
| Excercise | 2519 | 275 | 11% [10-12] | 278 | 11% [10-12] | 101 | 4% | 177 | 7% | 174 | 7% | 2067 | 82% |
| Colds | 2555 | 126 | 5% [4-6] | 335 | 13% [12-14] | 42 | 2% | 293 | 11% | 84 | 3% | 2136 | 84% |
| Pollen | 2517 | 58 | 2% [2-3] | 135 | 5% [5-6] | 26 | 1% | 109 | 4% | 32 | 1% | 2350 | 93% |
| Cold air | 2482 | 69 | 3% [2-4] | 65 | 3% [2-3] | 16 | 1% | 49 | 2% | 53 | 2% | 2364 | 95% |
| Pets | 2494 | 40 | 2% [1-2] | 58 | 2% [2-3] | 20 | 1% | 38 | 2% | 20 | 1% | 2416 | 97% |
| Laughter | 2481 | 61 | 2% [1-2] | 38 | 2% [2-3] | 10 | 0% | 28 | 1% | 51 | 2% | 2392 | 96% |
| House dust | 2501 | 54 | 2% [2-3] | 55 | 2% [2-3] | 14 | 1% | 41 | 2% | 40 | 2% | 2406 | 96% |

Wheeze, exertional wheeze and triggers of wheeze refer to the past 12 months. CI: confidence interval.

**Table S3:** Median difference in FeNO and FEV1/FVC and mean difference in FEV1 and FEF25-75 in schoolchildren with wheeze when reported by (A) children, (B) parents, and (C) parents and children, compared to children with no reported wheeze.

| | | FeNO, ppb [95% CI] | | | FEV1/FVC, % [95% CI] | | | FEV1, z-scores [95% CI] | | | FEF25-75, z-scores [95% CI] | | |
|---|---|---|---|---|---|---|---|---|---|---|---|---|---|
| | | Median | Crude diff.# | Adjusted diff.## | Median | Crude diff.# | Adjusted diff.## | Mean | Crude diff.+ | Adjusted diff.++ | Mean | Crude diff.+ | Adjusted diff.++ |
| **Wheeze** | | | | | | | | | | | | | |
| (A) | Children no | 11.6 | Ref. | Ref. | 86.9 | Ref. | Ref. | -0.527 | Ref. | Ref. | -0.554 | Ref. | Ref. |
| | Children yes | 19.3 | 7.7<br>[5.9, 9.5]<br>*** | 6.0<br>[4.4, 7.6]<br>*** | 83.9 | -3.1<br>[-4.3, -2.0]<br>*** | -3.2<br>[-4.4, -2.1]<br>*** | -0.649 | -0.122<br>[-0.254,0.011]<br>* | -0.101<br>[-0.232, 0.031] | -0.947 | -0.393<br>[-0.537,-0.250]<br>*** | -0.385<br>[-0.528, -0.242]<br>*** |
| (B) | Parents no | 11.6 | Ref. | Ref. | 86.9 | Ref. | Ref. | -0.521 | Ref. | Ref. | -0.557 | Ref. | Ref. |
| | Parents yes | 22.4 | 10.8<br>[8.8, 12.8]<br>*** | 11.2<br>[9.4, 13.0]<br>*** | 83.9 | -3.0<br>[-4.2, -1.8]<br>*** | -3.1<br>[-4.3, -1.9]<br>*** | -0.745 | -0.224<br>[-0.368,-0.080]<br>** | -0.200<br>[-0.342, -0.057]<br>** | -0.984 | -0.427<br>[-0.582,-0.273]<br>*** | -0.417<br>[-0.571, -0.263]<br>*** |
| (C) | Parents & children agreed in no | 11.4 | Ref. | Ref. | 87.0 | Ref. | Ref. | -0.519 | Ref. | Ref. | -0.540 | Ref. | Ref. |
| | Parents & children disagreed | 16.6 | 5.3<br>[3.5, 7.1]<br>*** | 4.1<br>[2.6, 5.6]<br>*** | 85.2 | -1.8<br>[-2.9, -0.7]<br>** | -2.0<br>[-3.1, -0.9]<br>*** | -0.610 | -0.090<br>[-0.222,0.042] | -0.084<br>[-0.215, 0.047] | -0.826 | -0.287<br>[-0.429,-0.144]<br>*** | -0.284<br>[-0.426, -0.142]<br>*** |
| | Parents & children agreed in yes | 31.2 | 20.5<br>[17.7, 23.3]<br>*** | 20.7<br>[18.2, 23.1]<br>*** | 82.4 | -4.6<br>[-6.3,-2.9]<br>*** | -4.5<br>[-6.2, -2.8]<br>*** | -0.795 | -0.275<br>[-0.476,-0.075]<br>** | -0.232<br>[-0.432, -0.033]<br>** | -1.134 | -0.594<br>[-0.810,-0.378]<br>*** | -0.579<br>[-0.792, -0.361]<br>*** |
| N | | | 2674 | | | 2147 | | | 2479 | | | 2147 | |
| **Exertional wheeze** | | | | | | | | | | | | | |
| (A) | Children no | 11.6 | Ref. | Ref. | 86.9 | Ref. | Ref. | -0.509 | Ref. | Ref. | -0.549 | Ref. | Ref. |
| | Children yes | 16.4 | 4.8<br>[3.3, 6.3]<br>*** | 3.2<br>[1.9, 4.5]<br>*** | 85.3 | -1.6<br>[-2.6, -0.6]<br>** | -1.8<br>[-2.8, -0.8]<br>*** | -0.718 | -0.208<br>[-0.324,-0.093]<br>*** | -0.162<br>[-0.278, -0.047]<br>** | -0.883 | -0.334<br>[-0.461,-0.207]<br>*** | -0.311<br>[-0.439, -0.184]<br>*** |
| (B) | Parents no | 11.6 | Ref. | Ref. | 86.8 | Ref. | Ref. | -0.515 | Ref. | Ref. | -0.560 | Ref. | Ref. |
| | Parents yes | 21.8 | 10.2<br>[8.2,12.2]<br>*** | 10.4<br>[8.7, 12.1]<br>*** | 84.2 | -2.6<br>[-3.8, -1.4]<br>*** | -3.1<br>[-4.3, -1.8]<br>*** | -0.776 | -0.261<br>[-0.403,-0.120]<br>*** | -0.201<br>[-0.342, -0.059]<br>** | -0.944 | -0.383<br>[-0.539,-0.228]<br>*** | -0.356<br>[-0.512, -0.200]<br>*** |
| (C) | Parents & children agreed in no | 11.5 | Ref. | Ref. | 86.9 | Ref. | Ref. | -0.502 | Ref. | Ref. | -0.542 | Ref. | Ref. |
| | Parents & children disagreed | 15.2 | 3.7<br>[2.0, 5.4]<br>*** | 2.0<br>[0.6, 3.5]<br>** | 86.1 | -0.8<br>[-1.8, 0.3] | -1.0<br>[-2.1, 0.0]<br>** | -0.662 | -0.160<br>[-0.281,-0.039]<br>** | -0.125<br>[-0.245, -0.004]<br>** | -0.752 | -0.211<br>[-0.345,-0.077]<br>** | -0.195<br>[-0.320, -0.061]<br>** |
| | Parents & children agreed in yes | 24.8 | 13.4<br>[10.7, 16.1]<br>*** | 13.3<br>[11.0, 15.6]<br>*** | 83.6 | -3.3<br>[-4.9, -1.8]<br>*** | -3.7<br>[-5.2, -2.1]<br>*** | -0.838 | -0.336<br>[-0.520,-0.153]<br>*** | -0.262<br>[-0.446, -0.078]<br>** | -1.090 | -0.549<br>[-0.748,-0.349]<br>*** | -0.515<br>[-0.715, -0.314]<br>*** |
| N | | | 2726 | | | 2189 | | | 2519 | | | 2189 | |

Wheeze and exertional wheeze refer to the past 12 months. #Unadjusted and ##adjusted by age and sex quantile regression models. +Unadjusted and ++ adjusted by age and sex linear regression models. P values *<0.1, ** <0.05, ***<0.001. CI: confidence interval. FeNO: Fractional exhaled nitric oxide. FEV1: forced expiratory volume in 1 second. FVC: forced vital capacity. FEV1/FVC: forced expiratory volume in 1 second over forced vital capacity. FEF25-75: forced expiratory flow between 25 and 75% of the forced vital capacity. Z-scores for FEV1 and FEF25-75 based on Global Lung Function Initiative references. Z-scores for FEV1 and FEF25-75 based on Global Lung Function Initiative references.

**Table S4:** Difference in median FeNO and FEV1/FVC and mean FEV1 and FEF25-75 in schoolchildren with wheeze between scenario A (child-reported) and B (parent-reported), scenario A and C (parent- and child-reported), and scenario B and C.

| | | FeNO, ppb [95% CI] | | | FEV1/FVC, % [95% CI] | | | FEV1, z-scores [95% CI] | | | FEF25-75, z-scores[95% CI] | | |
|---|---|---|---|---|---|---|---|---|---|---|---|---|---|
| | | Median | Crude diff.¶ | Adjusted diff. ¶¶ | Median | Crude diff. ¶ | Adjusted diff. ¶¶ | Mean | Crude diff. ǂ | Adjusted diff.ǂǂ | Mean | Crude diff. ǂ | Adjusted diff. ǂǂ |
| **Wheeze** | | | | | | | | | | | | | |
| (A - B) | Children yes | 19.3 | -3.1 [-8.7, 2.5] | -5.2 [-11.2, 0.7] * | 83.9 | 0.0 [-1.5, -1.5] | -0.1 [-1.7, 1.4] | -0.649 | 0.096 [-0.046, 0.239] | 0.099 [-0.065, 0.263] | -0.947 | 0.037 [-0.127, 0.201] | 0.032 [-0.157, 0.221] |
| | Parents yes | 22.4 | | | 83.9 | | | -0.745 | | | -0.984 | | |
| (A - C) | Children yes | 19.3 | -11.9 [-20.0, -3.8] ** | -14.7 [-21.7, -7.7] *** | 83.9 | 1.5 [0.0, 2.9] ** | 1.3 [-0.3, 2.8] | -0.649 | 0.146 [0.007, 0.284] ** | 0.132 [-0.005, 0.269] * | -0.947 | 0.186 [0.011, 0.362] ** | 0.192 [-0.005, 0.379] ** |
| | Parents & children yes | 31.2 | | | 82.4 | | | -0.795 | | | -1.134 | | |
| (B - C) | Parents yes | 22.4 | -8.8 [-16.5, -1.1] ** | -9.5 [-16.1, -2.9] ** | 83.9 | 1.5 [0.1, 2.9] ** | 1.4 [-0.2, 2.9] * | -0.745 | 0.049 [-0.088, 0.187] | 0.033 [-0.108, 0.173] | -0.984 | 0.149 [-0.016, 0.314] * | 0.160 [-0.015, 0.201] * |
| | Parents & children yes | 31.2 | | | 82.4 | | | -0.795 | | | -1.134 | | |
| **Exertional wheeze** | | | | | | | | | | | | | |
| (A - B) | Children yes | 16.4 | -5.4 [-9.8, -0.9] ** | -7.2 [-12.4, -2.0] ** | 85.3 | 1.1 [-0.5, 2.7] | 1.2 [-0.3, 2.8] | -0.718 | 0.058 [-0.070, 0.187] | 0.038 [-0.099, 0.175] | -0.883 | 0.061 [-0.085, 0.207] | 0.045 [-0.112, 0.202] |
| | Parents yes | 21.8 | | | 84.2 | | | -0.776 | | | -0.944 | | |
| (A - C) | Children yes | 16.4 | -8.4 [-14.6, -2.2] ** | -10.1 [-17.4, -2.9] ** | 85.3 | 1.7 [0.1, 3.4] ** | 1.9 [0.3, 3.4] ** | -0.718 | 0.120 [-0.028, 0.269] | 0.100 [-0.043, 0.242] | -0.883 | 0.207 [0.040, 0.375] ** | 0.203 [0.044, 0.363] ** |
| | Parents & children yes | 24.8 | | | 83.6 | | | -0.838 | | | -1.090 | | |
| (B - C) | Parents yes | 21.8 | -3.1 [-8.3, 2.2] | -2.9 [-8.6, 2.8] | 84.2 | 0.6 [-0.8, 2.1] | 0.6 [-0.8, 2.1] | -0.776 | 0.062 [-0.059, 0.183] | 0.061 [-0.053, 0.176] | -0.944 | 0.146 [0.009, 0.284] ** | 0.159 [0.019, 0.298] ** |
| | Parents & children yes | 24.8 | | | 83.6 | | | -0.838 | | | -1.090 | | |

Wheeze and exertional wheeze refer to the past 12 months. ¶ Unadjusted difference in medians and 95% confidence intervals (CI) calculated using the percentile bootstrap method with 500 repetitions. ¶¶ Difference in medians adjusted for age and sex with quantile regression models and 95% bootstrap CI. ǂUnadjusted difference in means and 95% bootstrap CI. ǂǂ Difference in means adjusted for age and sex with linear regression models and 95% bootstrap CI. P values *<0.1, ** <0.05, ***<0.001. FeNO: Fractional exhaled nitric oxide. FEV1: forced expiratory volume in 1 second. FVC: forced vital capacity. FEV1/FVC: forced expiratory volume in 1 second over forced vital capacity. FEF25-75: forced expiratory flow between 25 and 75% of the forced vital capacity. Z-scores for FEV1 and FEF25-75 based on Global Lung Function Initiative references.

**Table S5:** Difference in median FeNO and FEV1/FVC in schoolchildren with wheeze when reported by children compared to reported by parents stratified by age groups.

| | | FeNO, ppb [95% CI] | | | FEV1/FVC, % [95% CI] | | | FEV1 z-scores [95% CI] | | | FEV1 z-scores [95% CI] | | |
|---|---|---|---|---|---|---|---|---|---|---|---|---|---|
| | | Median | Crude diff.¶ | Adjusted diff. ¶¶ | Median | Crude diff. ¶ | Adjusted diff. ¶¶ | Mean | Crude diff. ‡ | Adjusted diff.‡‡ | Mean | Crude diff. ‡ | Adjusted diff. ‡‡ |
| **Wheeze** | (A - B) | | | | | | | | | | | | |
| <10 years | Children yes | 12.6 | 0.4 [-5.7, 6.4] | -0.3 [-7.3, 6.7] | 83.8 | 0.3 [-4.2, 4.9] | 0.5 [-3.7, 4.7] | -0.514 | 0.146 [-0.187, 0.479] | 0.149 [-0.190, 0.489] | -0.789 | 0.094 [-0.271, 0.458] | 0.102 [-0.284, 0.488] |
| | Parents yes | 12.2 | | | 83.5 | | | -0.684 | | | -0.882 | | |
| ≥ 10 years | Children yes | 21.0 | -7.7 [-14.6, -0.8] | -7.2 [-14.0, -0.4] | 83.9 | 0.0 [-1.7, 1.7] | -0.1 [-1.8, 1.7] | -0.684 | 0.082 [-0.086, 0.250] | 0.084 [-0.079, 0.247] | -0.993 | 0.019 [-0.170, 0.208] | 0.016 [-0.198, 0.230] |
| | Parents yes | 28.7 | ** | ** | 83.9 | | | -0.766 | | | -1.012 | | |
| **Exertional wheeze** | (A - B) | | | | | | | | | | | | |
| <10 years | Children yes | 12.1 | -5.8 [-15.0, 3.4] | -5.5 [-15.1, 4.1] | 85.8 | 3.6 [-1.4, 8.6] | 0.8 [-4.0, 5.7] | -0.407 | 0.084 [-0.246, 0.414] | 0.093 [-0.246, 0.433] | -0.715 | 0.141 [-0.227, 0.510] | 0.126 [-0.266, 0.518] |
| | Parents yes | 17.9 | | | 82.2 | | | -0.491 | | | -0.857 | | |
| ≥ 10 years | Children yes | 16.6 | -6.8 [-12.6, -0.9] | -6.9 [-13.1, -0.73] | 85.2 | 0.8 [-1.0, 2.6] | 0.8 [-0.8, 2.5] | -0.763 | 0.055 [-0.085, 0.194] | 0.037 [-0.120, 0.194] | -0.911 | 0.048 [-0.124, 0.219] | 0.041 [-0.142, 0.224] |
| | Parents yes | 23.4 | ** | ** | 84.4 | | | -0.818 | | | -0.959 | | |

Wheeze and exertional wheeze refer to the past 12 months. ¶ Unadjusted difference in medians and 95% confidence intervals (CI) calculated using the percentile bootstrap method with 500 repetitions. ¶¶ Difference in medians adjusted for age and sex with quantile regression models and 95% bootstrap CI. ‡Unadjusted difference in means and 95% bootstrap CI. ‡‡ Difference in means adjusted for age and sex with linear regression models and 95% bootstrap CI. P values *<0.1, ** <0.05, ***<0.001. FeNO: Fractional exhaled nitric oxide. FEV1: forced expiratory volume in 1 second. FVC: forced vital capacity. FEV1/FVC: forced expiratory volume in 1 second over forced vital capacity. FEF25-75: forced expiratory flow between 25 and 75% of the forced vital capacity. Z-scores for FEV1 and FEF25-75 based on Global Lung Function Initiative references.

**Figure S1:** Comparison of characteristics between parent-child agreement groups using multinomial logistic regression adjusted for all factors in the table.

**(1)Parents and children agreed for yes vs disagreed.** **(2)Parents and children agreed for no vs disagreed.**

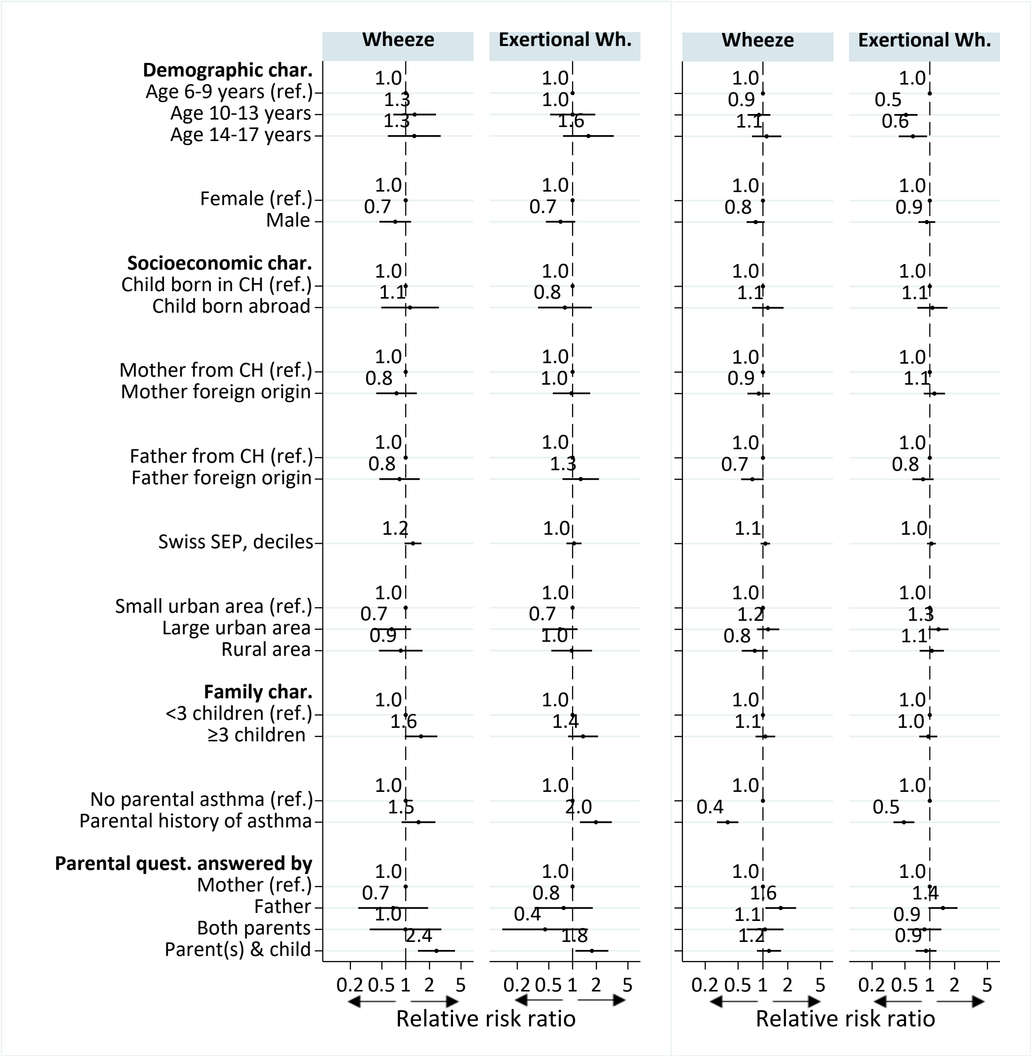


Interpretation of relative risk ratio:

Less likely to agree ← 1 → More likely to agree
Relative risk ratio

Parent-child agreement outcome groups:

| | | Parent-reported wheeze | |
|---|---|---|---|
| | | yes | no |
| Child-reported wheeze | yes | (1) Agreed for yes | (ref) Disagreed |
| | no | (ref) Disagreed | (2) Agreed for no |

Wheeze and exertional wheeze refer to the past 12 months. Abbreviations: Wh.=Wheeze; char.=characteristics; CH: Switzerland; quest.: questionnaire; Swiss SEP: Swiss socioeconomic position index. The Swiss SEP is an area-based measure developed by the Swiss Nation cohort that ranges from 0 to 100 and incorporates information on rent per square meter, education and occupation of household heads, and number of persons per household room. The urbanization degree was categorized according to the classification of the Swiss federal office of statistics classification. Graphs created using the STATA command coefplot [1].
